# Identification of Genetic Loci Simultaneously Associated with Multiple Cardiometabolic Traits

**DOI:** 10.1101/2021.12.13.21267561

**Authors:** Alexis C. Wood, Amit Arora, Michelle Newell, Victoria L. Bland, Jin Zhou, Nicola Pirastu, Jose M. Ordovas, Yann C. Klimentidis

## Abstract

**Background and Aims:** Cardiometabolic disorders (CMD) arise from a constellation of features such as increased adiposity, hyperlipidemia, hypertension and compromised glucose control. Many genetic loci have shown associations with individual CMD-related traits, but no investigations have focused on simultaneously identifying loci showing associations across all domains. We therefore sought to identify loci associated with risk across seven continuous CMD-related traits.

**Methods and Results:** We conducted separate genome-wide association studies (GWAS) for systolic and diastolic blood pressure (SBP/DBP), hemoglobin A1c (HbA1c), low- and high-density lipoprotein cholesterol (LDL-C/HDL-C), waist-to-hip-ratio (WHR), and triglycerides (TGs) in the UK Biobank (N=356,574–456,823). Multiple loci reached genome-wide levels of significance (N=145-333) for each trait, but only four loci (in/near *VEGFA, GRB14-COBLL1, KLF14*, and *RGS19-OPRL1)* were associated with risk across all seven traits (P<5×10^−8^). We sought replication of these four loci in an independent set of seven trait-specific GWAS meta-analyses. *GRB14-COBLL1* showed the most consistent replication, revealing nominally significant associations (P<0.05) with all traits except DBP.

**Conclusions:** Our analyses suggest that very few loci are associated in the same direction of risk with traits representing the full spectrum of CMD features. We identified four such loci, and an understanding of the pathways between these loci and CMD risk may eventually identify factors that can be used to identify pathologic disturbances that represent broadly beneficial therapeutic targets.

## Introduction

Cardiometabolic risk, the chance of developing type 2 diabetes (T2D), cardiovascular disease (CVD) and/or stroke, is typically assessed through measures of adiposity, glucose control, lipid metabolism, and blood pressure [1]. The clustering of these risk factors has been termed CMD and is defined by The American College of Cardiology as the presence of hypertension, poor glucose control, dyslipidemia, and abdominal obesity [2]. Recent data from the National Health and Nutrition Examination Survey (NHANES) indicate that among US adults, almost 90% of those with obesity, 50% with overweight and 25% of those at a normal weight met criteria for elevated risk in at least of these indicators [3]. While individual measures of cardiometabolic risk are linked with the development of CVD and T2D, studies are not unified in identifying which specific measures convey the greatest risk for these outcomes [4, 5]. Where evidence does strongly converge is in showing that the number of cardiometabolic risk factors is directly proportional to the risk of developing endpoints such as T2D, CVD, and for predicting mortality [6].

Each CMD feature results from a complex interplay between host genetics and lifestyle factors. Health behaviors such as physical activity and good dietary habits are associated with protection across multiple cardiometabolic risk factors [7], suggesting at least a partially shared behavioral etiology underlying CMD traits. However, the role that genetics plays in any shared etiology between CMD traits is not well understood[8, 9]. For example, only two loci have previously been identified as being implicated across T2D and cardiovascular disease: *CDKN2A/B* and *IRS1*[9, 10]. Identifying those genetic factors that convey risk across multiple CMD traits may increase our understanding of why risk factor clustering is not uniform across individuals, and further help identify those whose overall CMD profile suggests a greater risk of T2D and CVD and whom may especially benefit from behavioral interventions [11]. All CMD indicators show significant heritability [12, 13], with modest genetic correlations in the range of .1 -.5, suggesting both shared and measure-specific genetic loci across all traits associated with CMD [14, 15]. In addition, a small number of loci have been identified that increase the risk for one indicator (e.g., low-density lipoprotein cholesterol) but decrease the risk for another (e.g., glucose control [16–19]), mirroring some of the therapeutic effects of statins which reduce dyslipidemia but convey a slightly increased risk for T2D [20–22]. However, we are not aware of any studies that seek to identify variants associated with all the cardiometabolic risk factors forming the features of CMD. The identification of such loci, if they exist, may help identify underlying etiological pathways shared across many risk factors, improve our risk stratification efforts and provide information on targets for therapeutics which, by seeking to address multiple CMD risk factors, may have a broader range of beneficial clinical changes [23–25].

The present study aims to identify loci that are associated with each of seven cardiometabolic traits, representing the four features of CMD, in the same (risk predisposing) direction. Using data from the UK Biobank, which allowed us to leverage large numbers of individuals with both genetic data and phenotype data across multiple indicators of cardiometabolic disease, we took a genome-wide approach to identify variants significantly associated with each of glucose control, central adiposity, and multiple measures of lipid metabolism, and blood pressure. We then validated any significant associations in the published summary statistics from independent large-scale meta-analytic GWAS for each trait.

## Methods

### Study Population

The UK Biobank is a multi-center prospective cohort study of 500,000 adults ages 39 to 72 living in the UK. At each one of the 21 assessment centers, participants answered questionnaires, were interviewed, and were physically measured as part of a baseline visit that occurred between 2006 and 2010. A blood draw in a non-fasting state was performed for subsequent genetic and biomarker analysis. Among the biomarkers tested were directly-measured circulating LDL-C, HDL-C, hemoglobin A1c (HbA1c), TG, and alanine aminotransferase (ALT). The UK Biobank was approved by the National Information Governance Board for Health and Social Care and the National Health Service North West Multicentre Research Ethics Committee. All participants provided informed consent. Prevalent T2D was defined according to the following criteria: 1) self-reported T2D or generic diabetes in verbal interviews, 2) over the age of 35 years at diagnosis, and 3) not using insulin within one year of diagnosis, to exclude possible type-1 diabetes cases [26].

### Genotype data

DNA of participants was genotyped with the Affymetrix UK Biobank Axiom Array (Santa Clara, CA, USA), except for 10% of participants genotyped with the Affymetrix UK BiLEVE Axiom Array. Information about genotype imputation, principal components analysis, and quality control procedures can be found elsewhere [27]. Briefly, individuals were excluded if they exhibited unusually high heterozygosity, a high missing rate (>5%), or exhibited a mismatch between genetically inferred sex and self-reported sex. SNPs were excluded from analyses based on Hardy Weinberg equilibrium (p<1 ×10^−6^), high missing rate (>1.5%), low minor allele frequency (<0.1%), or a low imputation accuracy (info<0.4).

### GWAS and Genetic Correlations of Trait Measures

We first examined seven continuous cardiometabolic traits in the UK Biobank: HbA1c, LDL-C, HDL-C, TGs, waist-to-hip ratio adjusted for BMI (WHRadjBMI), and SBP and DBP adjusted for BMI. Although fasting glucose is a commonly used glycemic trait, since UKB participants are not fasted at the time of blood draw, we chose to use HbA1c instead. This allows us to achieve a similar sample size and power for each of the seven traits in the discovery analysis. Although HbA1c does have some limitations, is it more stable and reflective of long-term glycemia than fasting glucose. For the analyses of TG and HDL-C, we excluded individuals reporting cholesterol-lowering medications. For the analysis of LDL-C, we adjusted the values of people reporting cholesterol-lowering medication, by dividing their LDL-C values by a correction factor of 0.63. For the analysis of HbA1c, we excluded individuals with prevalent T2D (see above). For the analyses of SBP and DBP, we obtained the mean values across two manual measurements and adjusted for the effect of anti-hypertensive medications by adding 15 and 10 points to SBP and DBP, respectively, among individuals reported to be on anti-hypertensive medications (and an additional 2 measurements in a subset of individuals) [28]. We used LD-score regression to estimate genetic correlations across all pairs of trait [29].

### GWAS Replication Associations

We replicated identified associations by examining summary statistics of previously conducted large-scale GWAS of the same traits: LDL-C, HDL-C, and TG from the Global Lipids Genetics Consortium (GLGC; n=188,577)[30], WHRadjBMI from the Genetic Investigation of Anthropometric Traits (GIANT) consortium (n=224,459) [31], SBP and DBP from Genetic Epidemiology Research on Adult Health and Aging (GERA) cohort (n=99,785) [32], and HbA1c from the Meta-Analyses of Glucose and Insulin-related traits Consortium (MAGIC; n= 123,665) [33].

### Follow-up associations with CMD end points

We examined the association of the four identified SNPs with coronary artery disease (CAD), T2D, and stroke, some of the major outcomes arising from CMD, using GWAS meta-analysis results published by the Coronary Artery Disease Genome-wide Replication And Meta-analysis (CARDIOGRAM; n= up to 185,000)[34], Diabetes Genetics Replication And Meta-analysis (DIAGRAM; n= 898,130)[34], and MEGASTROKE (n= 521612) [35] consortia.

### Statistical analysis

All UK Biobank phenotypes were inverse-normalized prior to GWAS. Although this limits our ability to interpret effect sizes, it does allow us to both normalize the variables and to compare effect sizes across traits. GWAS was performed with BOLT-LMM software which implements a linear mixed model regression that includes a random effect consisting of all SNP genotypes other than the one being tested.[36] We included the following variables as covariates: age, age^2^, sex, center, genotyping chip, and the first 10 principal components. SNPs were selected if their beta coefficient had the same direction of effect (opposite direction for HDL-C) and a corresponding p-value<5 × 10^−8^ for each of the seven traits. If several SNPs were within a 1 Mb region, we chose the one with the lowest p-value based on a multivariate analysis conducted with metaCCA [37]. We also performed multiple-trait genetic colocalization using the “*hyprcoloc*” package in R to identify the best candidate variant given a genomic region for each locus, using a region ± 250kb region around the lead SNP.[38] A posterior probability of full colocalization (PPFC) threshold of 0.72 (e.g. alignment probability=0.85 and regional probability=0.85) was considered. In the replication dataset, we examined SNPs identified in the UK Biobank discovery set, and considered that a consistent direction of effect, and with p<0.05, qualified as a replication, given the smaller sample sizes in the replication datasets.

## Results

### UK Biobank GWAS

Phenotypic and genetic correlations among seven traits are shown in **Supplementary Figure 1**. Both phenotypic and genetic correlations were generally weak, with the exception of some correlations among some lipid traits and between SBP and DBP. In GWAS among UK Biobank participants, we identified 16 SNPs at four loci associated at genome-wide significance with each of seven quantitative traits in the same risk-conferring/protecting direction, in/near *VEGFA, GRB14-COBLL1, KLF14*, and *RGS19-OPRL1* (**Table 1 and Figure 1**). In sex-stratified analyses, there were no genome-wide significant SNPs for each trait (**Figure 1, Supplementary Table 1**). Colocalization analyses showed colocalization across all seven traits for the *GRB14*-*COBBL1* locus, across six traits for *KLF14* and *RGS19*-*OPRL1*, and across five traits for *VEGFA* (all PPFC>0.72; **Supplementary Table 2**).

**Table 1.** SNPs with significant associations in initial UK Biobank GWAS

|  | n | *GRB14-COBL1*<br>rs13389219:C/T |  |  | *KLF14*<br>rs201632637:CTTTTTT/C |  |  | *RGS19-OPRL1*<br>rs8126001: C/T |  |  | *VEGFA-C6orf223*<br>rs998584: C/A |  |  |
| --- | --- | --- | --- | --- | --- | --- | --- | --- | --- | --- | --- | --- | --- |
| | | $\beta$ | SE | p | $\beta$ | SE | p | $\beta$ | SE | p | $\beta$ | SE | p |
| TG (mmol/L) | 356,574 | 0.037 | 0.002 | 7.7E-63 | 0.032 | 0.002 | 1.8E-50 | 0.017 | 0.002 | 2.6E-15 | -0.040 | 0.002 | 2.6E-76 |
| LDL-C (mmom/L) | 431,167 | 0.018 | 0.002 | 2.4E-21 | 0.018 | 0.002 | 6.9E-22 | 0.017 | 0.002 | 1.0E-18 | -0.017 | 0.002 | 1.3E-18 |
| HDL-C (mmol/L) | 397,721 | -0.028 | 0.002 | 1.8E-50 | -0.031 | 0.002 | 5.8E-63 | -0.012 | 0.002 | 7.1E-11 | 0.035 | 0.002 | 3.2E-80 |
| HbA1c (mmol/mol) | 417,018 | 0.012 | 0.002 | 2.5E-09 | 0.011 | 0.002 | 2.6E-08 | 0.012 | 0.002 | 1.4E-09 | -0.020 | 0.002 | 2.8E-26 |
| SBP (mmHg) | 453,413 | 0.016 | 0.002 | 3.4E-16 | 0.015 | 0.002 | 8.3E-14 | 0.018 | 0.002 | 7.1E-19 | -0.011 | 0.002 | 2.4E-08 |
| DBP (mmHg) | 453,413 | 0.015 | 0.002 | 1.7E-14 | 0.017 | 0.002 | 3.8E-17 | 0.012 | 0.002 | 3.7E-10 | -0.016 | 0.002 | 1.1E-16 |
| WHRadjBMI | 456,823 | 0.026 | 0.002 | 3.4E-64 | 0.012 | 0.001 | 1.7E-15 | 0.013 | 0.001 | 3.1E-17 | -0.037 | 0.001 | 5.0E-139 |
| metacca-p |  |  |  | 5.4E-220 |  |  | 2.1E-149 |  |  | 1.0E-60 |  |  | 0.0E+00 |

Abbreviations: DBP: diastolic blood pressure; HDL-C: high-density lipoprotein cholesterol; LDL-C: low-density lipoprotein cholesterol; SBP: systolic blood pressure; TG: triglycerides; WHRadjBMI: waist-to-hip ratio adjusted for BMI

**Table 2.** Published associations for significant SNPs in GWAS discovery

|  | Sample size | *GRB14-COBL1*<br>rs13389219:C/T |  |  | *KLF14*<br>rs11765979:A/C |  |  | *RGS19-OPRL1*<br>rs7271530: C/T |  |  | *VEGFA-C6orf223*<br>rs998584: C/A |  |  |
| --- | --- | --- | --- | --- | --- | --- | --- | --- | --- | --- | --- | --- | --- |
|  |  | beta | se | p | beta | se | p | beta | se | p | beta | se | p |
| TG (mg/dl) | n<sub>max</sub>=177,782 | 0.0271 | 0.0034 | 2.60E-15 | 0.0247 | 0.0047 | 2.32E-07 | 0.009 | 0.0056 | 4.71E-02 | 0.0293 | 0.0037 | 3.42E-15 |
| LDL-C (mg/dl) | n<sub>max</sub>=173,012 | 0.022 | 0.0037 | 9.89E-08 | 0.0111 | 0.0052 | 3.26E-02 | 0.0093 | 0.0064 | 2.31E-02 | 0.0005 | 0.004 | 0.9359 |
| HDL-C (mg/dl) | n<sub>max</sub>=173,012 | -0.0226 | 0.0035 | 4.54E-10 | -0.0412 | 0.0048 | 3.11E-17 | -0.0101 | 0.0059 | 9.65E-02 | -0.026 | 0.0038 | 2.27E-11 |
| HbA1C (%) | n=123,665 | 0.0062 | 0.0018 | 4.28E-04 | 0.0033 | 0.0018 | 6.08E-02 | 0.0051 | 0.0019 | 6.21E-03 | -0.0083 | 0.0024 | 4.92E-04 |
| SBP (mmHg) | n=99,785 | 0.122 | NA | 0.031 | 0.0863 | NA | 0.11 | 0.199 | NA | 7.70E-03 | 0.085 | NA | 0.23 |
| DBP (mmHg) | n=99,785 | 0.0634 | NA | 0.085 | 0.0248 | NA | 0.48 | 0.142 | NA | 3.50E-03 | 0.0707 | NA | 0.12 |
| WHRadjBMI | n=694,649 | 0.027 | 0.0035 | 3.20E-15 | 0.0088 | 0.0043 | 0.041 | 0.012 | 0.0045 | 7.70E-03 | 0.043 | 0.0038 | 1.10E-29 |

Note: Nominally significant associations (P<.05) in bold, standard errors (SE) not available in the published summary statistics for blood pressure.
rs11765979 at *KLF14* was used as a proxy for rs201632637 ( $r^2=0.94$ , CEU)
Abbreviations: DBP: diastolic blood pressure; HDL-C: high-density lipoprotein cholesterol; LDL-C: low-density lipoprotein cholesterol; SBP: systolic blood
pressure; TG: triglycerides; HHRadjBMI: waist-to-hip ratio adjusted for BMI

**Figure 1:**
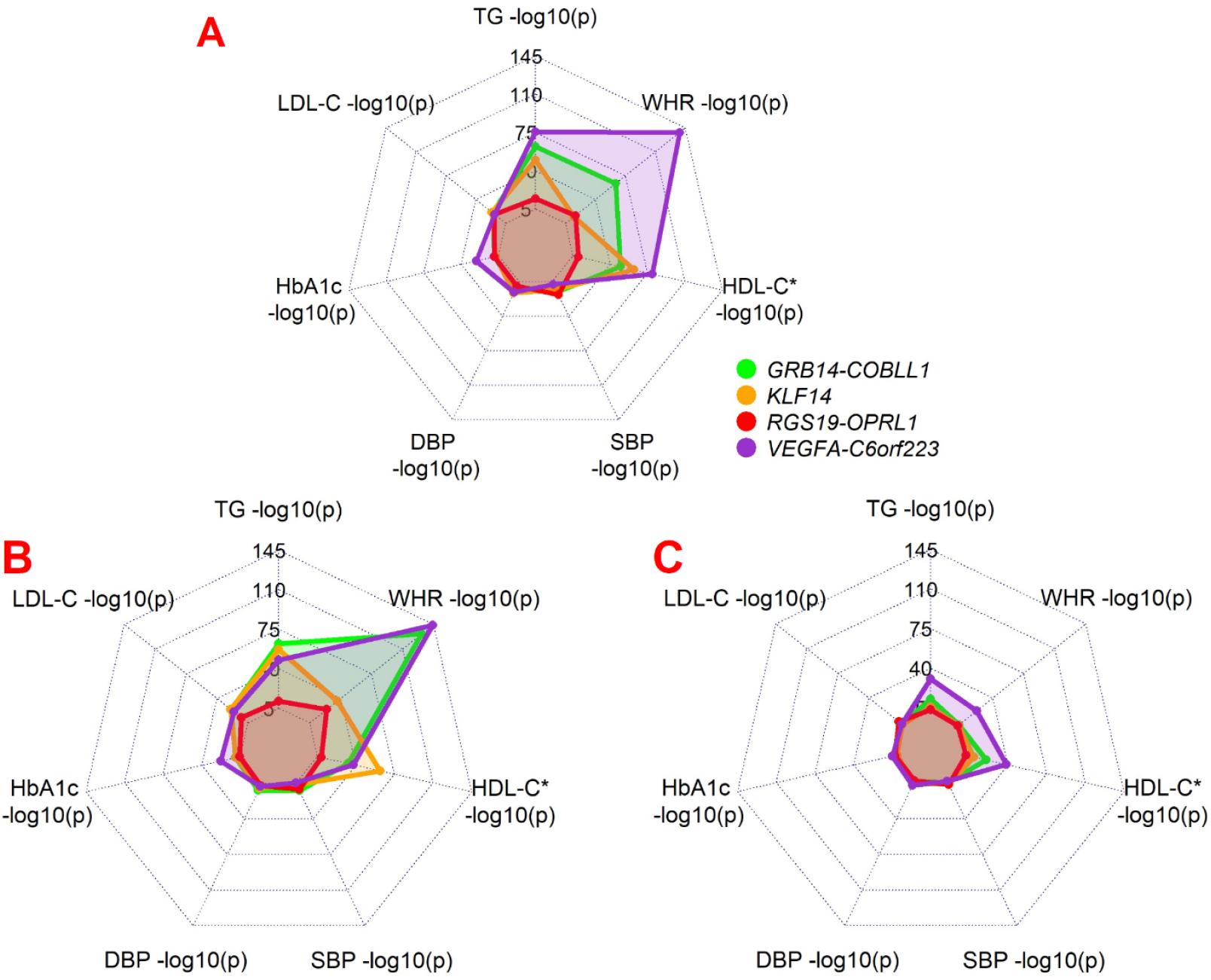
Radar plot showing association p-values for all four loci identified in UK Biobank in A) both men and women, B) women only, and C) men only.

### GWAS in replication samples

Among the four SNPs identified in UK Biobank, we found that all four exhibit the same direction of risk effect across all seven traits, but do not reach p<0.05 across all traits, likely due to the substantially smaller sample sizes in those studies as compared to the UK Biobank (**Table 2**).

### Follow-up associations with CMD end points and phenome-wide association study

We found directionally consistent associations of the four loci identified in UK Biobank with T2D, CAD, and stroke, although not all associations achieved statistical significance, particularly for stroke (**Supplementary Table 3**). In silico phenome-wide association analyses (PheWAS) of these four loci revealed associations with fat distribution patterns and cardiometabolic conditions and medications, with limited to no evidence of adverse associations with other conditions (see **Supplementary Tables 4-8**).

## Discussion

This is the first study, of which we are aware, to take a broad and well-powered genome-wide approach to simultaneously identifying genetic loci associated with seven cardiometabolic risk factors representing the four domains of CMD. We observed only four loci that increase the risk for all seven of our traits; raised TGs and LDL-C, lowered HDL-C, raised HbA1c, and systolic and diastolic blood pressure. Given that multiple robust genetic associations have been published for each of our seven traits individually, these findings suggest that loci which influence all seven features CMD are less common, and much of the genetic etiology is trait-specific.

We found four loci which simultaneously associated with risk across seven CMD traits at genome-wide levels of significance. Although this is the first study, of which we are aware to conduct discovery across all seven traits simultaneously, previous studies have identified associations between loci in *GRB14-COBLL1*, and *RGS19*-*OPRLI* with each of our cardiometabolic traits independently. The intergenic SNP rs13889216 in *GRB14-COBLL1*, or the nearby SNP rs10195252 in *GRB14* have previously been associated at genome-wide levels of significance with WHR ratio (sometimes with a larger effect size in females) [31, 39, 40] and levels of TGs [40–43], HDL-C [31, 40, 42–441] and LDL-C [31]. Other studies have associated these SNPs with additional traits often used as indicators of cardiometabolic disease or cardiometabolic disease risk, such as total cholesterol [43], T2D [31, 43, 45] and other indices of glycemic control [31, 40, 41, 43], often at levels approaching genome-wide significance. Similarly, rs8126001 found in the 5’ UTR of *OPRL1* and 2kb upstream from *RGS19* has been previously associated with TG levels [42], HDL-C [42], SBP[46] and WHRadjBMI as well as waist circumference in UK Biobank [46, 47], although its most replicated associations to date have been with alcohol use/abuse. Similarly, associations have been observed between variants at *KLF14* and *VEGFA* with both T2D [48] and HDL-C levels in large-scale GWAS [49]. Taken together, these previous data suggest our analysis approach is robust, and supporting the notion that our associations of each of these four loci with all seven traits is generalizable. In the light of evidence showing that the number of cardiometabolic risk factors is directly proportional to the risk of T2D, CVD and mortality [6], information that these loci associate with risk across multiple traits could be seen to raise the importance of these loci to the risk of developing these conditions when compared to loci which raise the risk profile of a fewer number of traits.

Understanding the mechanisms that link our four loci to CMD risk may yield insights into the full clinical profile of CMD. In the broader literature, two of our loci show multiple associations with mechanisms underlying, and/or patterns of, fat deposition. For example, *KLF14* regulates the expression of several genes in adipose tissue, and has been associated with favorable adiposity [50], and, in women with adipogenesis, shifts in central vs. gynoid adiposity, and the shift in the number and size of adipocytes [51]. In turn, *GRB14* and the *GRB14-COBLL1* locus have both been associated with body fat distribution (reviewed in [52]), and variation in or near *GRB14-COBLL1* to alter gene expression in subcutaneous fat, and in omental but not mesenteric visceral fat [53]. These findings suggest that at least some of our putative loci are united by their associations with patterns of fat deposition – an unsurprising implication given that fat deposition patterning has recently emerged as a stronger indicator of cardiometabolic risk factor clustering than overall adiposity, particularly in women (e.g. [54, 55]). In both genders, visceral fat is an indicator of increased cardiometabolic risk [54, 56–62], however in women only, deposition of fat in the lower body (gluteal-femoral fat) is associated with protection from CMD risk factors [59, 63] and their downstream endpoint of CVD [55]. Supporting the notion that CMD risk alleles may operate through fat patterning, a genetic risk score associated with fat deposition phenotypes such as WHR, gluteofemoral fat and abdominal fat volume, also associated CMD traits, as well as T2D and CVD [64], and the protective associations of *GRB14-COBLL1* variants on adverse lipid profiles have been theorized to me mediated through alterations in body fat distribution [52].

There are of course limitations to our findings. Our sample is comprised of individuals with European-ancestry and generalizations beyond this population are not advised. In addition, while the rationale for the current analyses was to identify loci which most increase the risk of cardiometabolic disease by exerting effects across multiple risk indicators simultaneously, our loci have not been widely associated with hard outcomes such as CVD, except for associations of *GRB14-COBLL1, KLF14*, and *VEGFA* with T2D (**Supplementary Table 4**) [31, 43, 45]. Finally, many of our loci have been observed to have sex-specific effects especially with regards to associations with fat patterning [51, 53, 65], and our analyses suggested sex-specific magnitudes of associations with the our seven CMD traits, we were not powered to detect these through formal interaction analyses. Future research should address these limitations, and also continue to examine the excess of risk associated with visceral fat and identify mechanisms linking fat deposition and cardiometabolic risk.

These current findings suggest that from all loci known to affect CMD related traits, only a few may have effects across multiple traits, which are strong enough to be readily detectable at genome-wide levels of significance. Although the effect sizes of these naturally occurring genetic variants are very small, they do not necessarily reflect the importance of the pathways that they point to. Interventions may thus have a much larger, more clinically meaningful impact than immediately apparent from the effect sizes of the SNP-risk factor associations used to identify these pathways. This has been illustrated in the case of *HMGCR* and statins with LDL-C [66, 67]. If these loci serve as an indicator of risk across the highest number of CMD features, and if future research identifies pathways linking these loci to CMD, such as via changes in fat patterning, this research may eventually give insights into therapeutic targets that will yield a broader range of clinical improvement.

## Data Availability

All data produced in the present study will be made available through the GWAS Catalog.

## Abbreviations

ALT: alanine aminotransferase
BMI: body mass index
CARDIOGRAM: Coronary Artery Disease Genome-wide Replication And Meta-analysis
CAD: coronary artery disease
CMD: cardiometabolic disorders
CVD: cardiovascular disease
DIAGRAM: Diabetes Genetics Replication And Meta-analysis
DBP: diastolic blood pressure
GERA: Genetic Epidemiology Research on Adult Health and Aging cohort
GIANT: Genetic Investigation of Anthropometric Traits consortium
GLGC: Global Lipids Genetics Consortium
GWAS: genome-wide association studies
HbA1c: hemoglobin A1c
HDL-C: high-density lipoprotein cholesterol
LDL-C: low-density lipoprotein cholesterol
NHANES: National Health and Nutrition Examination Survey
PPFC: posterior probability of full colocalization
SBP: systolic blood pressure
T2D: type 2 diabetes
TGs: triglycerides
UK: United Kingdom
WHR: waist-to-hip-ratio
WHRadjBMI: waist-to-hip ratio adjusted for BMI

## Acknowledgments

This research was conducted using the UK Biobank Resource under Application Number 15678. We thank the participants and organizers of the UK Biobank. The authors would also like to acknowledge the vital contributions of the GWAS consortia: GLGC, MAGIC, GIANT, DIAGRAM, CARDIOGRAM, as well as all organizers and participants of individual participating studies.

## Disclosures

The authors have no relevant conflicts of interest to disclose.

## Notes

**Sources of Support** The authors would like to acknowledge support from the National Heart, Lung, and Blood Institutes (R01-HL136528). Dr. Wood’s role on this project was funded, in part, by USDA/ARS cooperative agreement # 58-3092-5-001. The contents of this publication do not necessarily reflect the views or policies of the U.S. Department of Agriculture, nor does mention of trade names, commercial products, or organizations imply endorsement by the U.S. Government. The funders had no role in study design, data collection and analysis, decision to publish, or preparation of the manuscript.

### Competing Interest Statement

The authors have declared no competing interest.

### Funding Statement

This study was funded by the National Heart, Lung, and Blood Institutes (R01-HL136528), and the USDA/ARS cooperative agreement # 58-3092-5-001.

### Author Declarations

This study involves only open available data, which can be obtained from the UK Biobank and respective genetic consortia.

