## Supplementary for "Identification of Genetic Loci Simultaneously Associated with Multiple Cardiometabolic Traits"

**Supplementary Table 1:** Sex-specific associations of four loci identified in sex-combined analysis.

| | GRB14-COBLL1 rs13389219:C/T | | | KLF14 rs201632637:CTTTTTT/C | | | RGS19-OPRL1 rs8126001: C/T | | | VEGFA-C6orf223 rs998584: C/A | | |
|---|---|---|---|---|---|---|---|---|---|---|---|---|
| **Women** | beta | se | p | beta | se | p | beta | se | p | beta | se | p |
| TG (mmol/L) | 0.049 | 0.003 | 7.40E-63 | 0.046 | 0.003 | 9.60E-58 | 0.02 | 0.003 | 4.80E-12 | -0.042 | 0.003 | 1.90E-48 |
| LDL-C (mmom/L) | 0.026 | 0.003 | 1.10E-24 | 0.026 | 0.003 | 1.70E-25 | 0.019 | 0.003 | 2.70E-13 | -0.024 | 0.003 | 1.10E-21 |
| HDL-C (mmol/L) | -0.034 | 0.003 | 1.50E-33 | -0.047 | 0.003 | 1.00E-62 | -0.017 | 0.003 | 2.70E-09 | 0.036 | 0.003 | 1.90E-38 |
| HbA1c (mmol/mol) | 0.016 | 0.003 | 6.50E-10 | 0.016 | 0.003 | 1.10E-09 | 0.012 | 0.003 | 2.50E-06 | -0.026 | 0.003 | 1.50E-23 |
| SBP (mmHg) | 0.02 | 0.003 | 6.00E-13 | 0.014 | 0.003 | 2.80E-07 | 0.019 | 0.003 | 7.70E-12 | -0.012 | 0.003 | 2.10E-05 |
| DBP (mmHg) | 0.02 | 0.003 | 1.20E-12 | 0.017 | 0.003 | 1.30E-10 | 0.015 | 0.003 | 4.30E-08 | -0.016 | 0.003 | 5.80E-09 |
| WHRadjBMI | 0.067 | 0.003 | 2.60E-132 | 0.034 | 0.003 | 3.90E-37 | 0.028 | 0.003 | 4.80E-25 | -0.069 | 0.003 | 5.40E-145 |
| metacca-p | | | 0.00E+00 | | | 5.10E-154 | | | 1.20E-45 | | | 0.00E+00 |
| **Men** | | | | | | | | | | | | |
| TG (mmol/L) | 0.026 | 0.004 | 1.40E-13 | 0.017 | 0.003 | 1.80E-06 | 0.012 | 0.003 | 5.40E-04 | -0.041 | 0.003 | 3.40E-31 |
| LDL-C (mmom/L) | 0.009 | 0.003 | 1.60E-03 | 0.008 | 0.003 | 4.80E-03 | 0.015 | 0.003 | 7.10E-07 | -0.01 | 0.003 | 1.10E-03 |
| HDL-C (mmol/L) | -0.029 | 0.003 | 9.50E-21 | -0.019 | 0.003 | 1.20E-09 | -0.008 | 0.003 | 1.30E-02 | 0.04 | 0.003 | 3.30E-39 |
| HbA1c (mmol/mol) | 0.007 | 0.003 | 1.60E-02 | 0.004 | 0.003 | 1.90E-01 | 0.009 | 0.003 | 1.90E-03 | -0.013 | 0.003 | 1.60E-05 |
| SBP (mmHg) | 0.013 | 0.003 | 2.70E-05 | 0.015 | 0.003 | 7.40E-07 | 0.015 | 0.003 | 3.50E-07 | -0.011 | 0.003 | 3.60E-04 |
| DBP (mmHg) | 0.011 | 0.003 | 5.10E-04 | 0.015 | 0.003 | 1.00E-06 | 0.009 | 0.003 | 2.10E-03 | -0.017 | 0.003 | 3.90E-08 |
| WHRadjBMI | -0.008 | 0.003 | 1.30E-02 | -0.006 | 0.003 | 3.90E-02 | 0.003 | 0.003 | 2.80E-01 | -0.03 | 0.003 | 1.90E-22 |
| metacca-p | | | 1.40E-32 | | | 4.90E-18 | | | 3.70E-12 | | | 5.40E-79 |

**Supplementary Table 2:** Colocalization analyses at four top loci at PPFC>0.72 threshold

| Gene | Trait | N.snps | Posterior prob. | Regional prob. | Candidate SNP | Candidate SNP bp | allele1 | allele0 | Posterior explained by SNP |
|---|---|---|---|---|---|---|---|---|---|
| *KLF14* | TG, LDL, HDL, HBA1C, DBP, WHR | 2538 | 0.8869 | 0.9998 | 7:130438531_CTTTTTT_C | 130438531 | CTTTTTT | C | 0.9987 |
| *GRB14-COBBL1* | TG, LDL, HDL, HBA1C, SBP, DBP, WHR | 2888 | 0.9774 | 1 | rs13389219 | 165528876 | C | T | 0.9274 |
| *RSG19-OPRL1* | TG, LDL, HDL, HBA1C, SBP, WHR | 3405 | 0.9937 | 1 | rs8126001 | 62711459 | C | T | 1 |
| *VEGFA* | TG, LDL, HDL, HBA1C, WHR | 3575 | 0.998 | 1 | rs998584 | 43757896 | C | A | 1 |

**Supplementary Table 3:** Associations of top four loci with T2D (DIAGRAM), CAD (CARDIOGRAM), and stroke (METASTROKE)

| | ***GRB14-COBLL1*** rs13389219:C/T | | | ***KLF14*** rs11765979:A/C | | | ***RGS19-OPRL1*** rs8126001: C/T | | | ***VEGFA-C6orf223*** rs998584: C/A | | |
|---|---|---|---|---|---|---|---|---|---|---|---|---|
| | beta | se | p | beta | se | p | beta | se | p | beta | se | p |
| DIAGRAM | 0.0800 | 0.0076 | 8.60E-26 | 0.0580 | 0.0075 | 9.40E-15 | 0.0360 | 0.0076 | 2.30E-06 | -0.0550 | 0.0075 | 2.20E-13 |
| CARDIOGRAM | 0.0238 | 0.0101 | 1.88E-02 | 0.0192 | 0.0093 | 3.92E-02 | 0.0247 | 0.0108 | 2.29E-02 | -0.0419 | 0.0099 | 2.21E-05 |
| MEGASTROKE* | 0.014 | 0.0094 | 0.1393 | 0.0146 | 0.0109 | 0.1803 | 0.0139 | 0.0111 | 0.2119 | -0.012 | 0.0098 | 0.2208 |

*KLF14 used rs7271530: C/T

**Supplementary Table 4:** Phenome-wide association study of rs13389219 at COBLL1

| **Oxford Brain Imaging Genetics (BIG)** | | |
|---|---|---|
| **rs13389219 (Nearest genes: GRB14, COBLL1)** | | |
| **Phenotype** | **P-value** | **Beta** |
| Trunk fat percentage | 3.00E-27 | 0.025 |
| Hip circumference | 1.10E-26 | 0.026 |
| Body fat percentage | 1.90E-24 | 0.019 |
| Arm fat percentage (left) | 5.20E-20 | 0.017 |
| Arm fat percentage (right) | 1.20E-19 | 0.017 |
| Trunk fat mass | 2.40E-18 | 0.022 |
| Leg fat percentage (right) | 1.20E-16 | 0.013 |
| Impedance of arm (left) | 4.60E-16 | 0.014 |
| Impedance of arm (right) | 2.10E-15 | 0.014 |
| Whole body fat mass | 3.80E-15 | 0.019 |
| Waist-to-hip ratio adjusted for smoking | 1.10E-14 | 0.029 |
| Leg fat percentage (left) | 1.20E-14 | 0.012 |
| Leg fat mass (right) | 1.30E-12 | 0.014 |
| Arm fat mass (right) | 7.90E-12 | 0.017 |
| Waist-to-hip ratio adjusted for BMI | 8.50E-12 | 0.029 |
| Leg fat mass (left) | 1.00E-11 | 0.013 |
| Arm fat mass (left) | 6.80E-11 | 0.016 |
| Diabetes diagnosed by doctor | 1.90E-10 | -0.0034 |
| Impedance of whole body | 2.80E-10 | 0.012 |
| Treatment/medication code: metformin | 1.50E-07 | -0.002 |
| Non-cancer illness code, self-reported: diabetes | 4.60E-07 | -0.0024 |
| Weight | 7.20E-07 | 0.011 |
| Illnesses of siblings: Diabetes | 1.30E-06 | -0.0037 |
| Body mass index (BMI) | 2.20E-06 | 0.012 |
| Illnesses of mother: Diabetes | 2.60E-06 | -0.0035 |
| Non-cancer illness code, self-reported: type 2 diabetes | 1.20E-05 | -0.00087 |
| Medication for cholesterol, blood pressure, diabetes, or take exogenous hormones: Blood pressure medication | 2.60E-05 | -0.0054 |
| Non-cancer illness code, self-reported: hypertension | 4.60E-05 | -0.0044 |
| Treatment/medication code: latanoprost | 4.80E-05 | -0.00046 |
| Vascular/heart problems diagnosed by doctor: High blood pressure | 5.20E-05 | -0.0045 |
| Alcohol intake frequency. | 7.20E-05 | -0.014 |
| Medication for cholesterol, blood pressure, diabetes, or take exogenous hormones: None of the above | 1.80E-04 | 0.0059 |
| Vascular/heart problems diagnosed by doctor: None of the above | 2.40E-04 | 0.0042 |
| Treatment/medication code: zimovane ls 3.75mg tablet | 3.70E-04 | -0.00016 |
| Nap during day | 4.10E-04 | -0.0052 |
| Waist circumference adjusted for smoking | 4.10E-04 | 0.013 |
| Systolic blood pressure, automated reading | 4.40E-04 | -0.0089 |
| Illnesses of father: Diabetes | 5.60E-04 | -0.0027 |
| Total cholesterol in HDL | 6.60E-04 | 0.036 |
| Bipolar disorder status | 6.80E-04 | -0.079 |
| Trunk fat-free mass | 6.80E-04 | -0.0053 |
| Concentration of small VLDL particles | 7.20E-04 | -0.037 |
| Trunk predicted mass | 7.50E-04 | -0.0053 |
| Hand grip strength (left) | 7.90E-04 | -0.006 |
| Phospholipids in small VLDL | 7.90E-04 | -0.035 |
| Triglycerides in small VLDL | 8.00E-04 | -0.035 |
| Alcohol intake versus 10 years previously | 9.70E-04 | -0.0062 |
| Number of full sisters | 1.00E-03 | -0.0063 |
| Total lipids in small VLDL | 1.00E-03 | -0.036 |
| NETMAT amplitudes (100) 43 | 1.30E-03 | -0.046 |

| **Open Target Genetics** | | |
|---|---|---|
| **rs13389219 (Nearest gene: GRB14, COBLL1))** | | |
| **Trait** | **P-value** | **Beta** |
| Hip circumference | 1.69638E-31 | 0.257151 |
| Trunk fat percentage | 9.8901E-28 | 0.190711 |
| Body fat percentage | 8.45631E-26 | 0.160632 |
| Arm fat percentage (left) | 2.59868E-23 | 0.181244 |
| Arm fat percentage (right) | 3.16422E-23 | 0.179295 |
| Trunk fat mass | 1.51378E-22 | 0.121835 |
| Whole body fat mass | 1.92523E-20 | 0.207272 |
| High light scatter reticulocyte count | 2.54578E-20 | -0.000232181 |
| Leg fat percentage (right) | 1.468E-18 | 0.117006 |
| Leg fat mass (right) | 2.72291E-16 | 0.0310153 |
| Leg fat percentage (left) | 3.98294E-16 | 0.102969 |
| Arm fat mass (right) | 1.17982E-15 | 0.0119239 |
| Arm fat mass (left) | 1.65881E-15 | 0.0132548 |
| Impedance of arm (left) | 1.73805E-15 | 0.753797 |
| Triglycerides | 2.598E-15 | -0.0271 |
| Impedance of arm (right) | 2.66753E-15 | 0.729642 |
| Leg fat mass (left) | 2.70077E-15 | 0.02923 |
| Reticulocyte count | 1.36134E-14 | -0.000739579 |
| Diabetes mellitus | 3.36E-14 | -0.0834 |
| Reticulocyte count | 6.503E-14 | -0.02749336 |
| Type 2 diabetes | 3.8E-13 | -0.0826 |
| Reticulocyte percentage | 9.38069E-12 | -0.0152436 |
| High light scatter reticulocyte count | 1.113E-11 | -0.02485962 |
| Reticulocyte fraction of red cells | 1.145E-11 | -0.02487405 |
| Diabetes diagnosed by doctor | 2.18626E-11 | -0.075061017 |
| Haemoglobin concentration | 3.08655E-11 | -0.0158461 |
| Red blood cell (erythrocyte) count | 1.34624E-10 | -0.0054783 |
| Impedance of whole body | 1.37837E-10 | 1.03526 |
| High light scatter reticulocyte percentage of red cells | 4.43E-10 | -0.02283534 |
| Body mass index (bmi) | 2.43204E-09 | 0.0680645 |
| Body mass index (bmi) | 3.51935E-09 | 0.0676235 |
| High light scatter reticulocyte percentage | 3.91605E-09 | -0.00491419 |
| Weight | 2.88166E-08 | 0.188401 |
| Weight | 3.35496E-08 | 0.188249 |
| Metformin \| treatment/medication code | 6.3622E-08 | -0.083430798 |
| Mean sphered cell volume | 9.66984E-08 | 0.0688302 |
| LDL cholesterol | 9.894E-08 | -0.022 |
| Diabetes \| illnesses of siblings | 1.23286E-07 | -0.051000667 |
| Diabetes \| non-cancer illness code, self-reported | 1.33489E-07 | -0.065155238 |
| Haematocrit percentage | 1.47043E-07 | -0.0369737 |
| Essential hypertension | 0.000000238 | -0.0339 |
| Hemoglobin concentration | 2.642E-07 | -0.01863582 |
| Hypertension | 0.000000313 | -0.0335 |
| Type 2 diabetes | 0.00000079 | -0.076961041 |
| Cholesterol, total | 0.000001335 | -0.0199 |
| Mean reticulocyte volume | 1.46612E-06 | 0.0918663 |
| Lymphocyte percentage | 0.000003237 | -0.0824311 |
| Diabetes \| illnesses of mother | 3.2768E-06 | -0.039706024 |
| Alcohol intake frequency. | 3.97088E-06 | -0.0162193 |
| Blood pressure medication \| medication for cholesterol, blood pressure, diabetes, or tak | 4.7987E-06 | -0.038441701 |

**Supplementary Table 5:** Phenome-wide association study of rs7633256 at *KLF14*

| Oxford Brain Imaging Genetics (BIG) | | |
|---|---|---|
| **rs201632637 (proxy used: rs7633256, R2 = 0.94, Ref = CEU) (Nearest gene: KLF14)** | | |
| **Phenotype** | **P-value** | **Beta** |
| Hip circumference | 1.90E-16 | 0.02 |
| Trunk fat percentage | 6.00E-14 | 0.017 |
| Medication for cholesterol, blood pressure, diabetes, or take exogenous hormones: Blood pressure medication | 1.50E-13 | -0.0094 |
| Body fat percentage | 1.90E-13 | 0.014 |
| Medication for cholesterol, blood pressure, diabetes, or take exogenous hormones: Cholesterol lowering medication | 7.10E-11 | -0.0073 |
| Non-cancer illness code, self-reported: hypertension | 9.10E-11 | -0.007 |
| Vascular/heart problems diagnosed by doctor: High blood pressure | 1.30E-10 | -0.007 |
| Arm fat percentage (left) | 1.50E-10 | 0.012 |
| Vascular/heart problems diagnosed by doctor: None of the above | 2.60E-10 | 0.0071 |
| Arm fat percentage (right) | 6.60E-10 | 0.012 |
| Leg fat percentage (right) | 7.80E-10 | 0.0096 |
| Impedance of arm (right) | 8.10E-10 | 0.011 |
| Impedance of arm (left) | 8.70E-10 | 0.011 |
| Trunk fat mass | 1.10E-09 | 0.015 |
| Leg fat percentage (left) | 5.30E-09 | 0.009 |
| Medication for cholesterol, blood pressure, diabetes, or take exogenous hormones: None of the above | 6.00E-08 | 0.0085 |
| Whole body fat mass | 1.30E-07 | 0.013 |
| Impedance of whole body | 2.90E-07 | 0.0096 |
| Diabetes diagnosed by doctor | 8.80E-07 | -0.0026 |
| Treatment/medication code: simvastatin | 3.80E-06 | -0.0036 |
| Arm fat mass (left) | 8.20E-06 | 0.011 |
| Leg fat mass (right) | 1.40E-05 | 0.0085 |
| Arm fat mass (right) | 2.00E-05 | 0.01 |
| Non-cancer illness code, self-reported: high cholesterol | 2.00E-05 | -0.0034 |
| Leg fat mass (left) | 2.20E-05 | 0.0082 |
| Treatment/medication code: metformin | 2.20E-05 | -0.0016 |
| Treatment/medication code: lisinopril | 2.30E-05 | -0.0017 |
| Illnesses of mother: Diabetes | 3.00E-05 | -0.0031 |
| Systolic blood pressure, automated reading | 5.20E-05 | -0.01 |
| Non-cancer illness code, self-reported: diabetes | 7.80E-05 | -0.0019 |
| Comparative height size at age 10 | 1.10E-04 | 0.0065 |
| Age when periods started (menarche) | 1.50E-04 | -0.0095 |
| FreeSurfer a2009s rh S occipital ant thickness | 2.10E-04 | -0.055 |
| FreeSurfer DKTatlas rh parsorbitalis area | 2.50E-04 | -0.048 |
| Diagnoses - main ICD10: Z41 Procedures for purposes other than remedying health state | 3.00E-04 | 0.00032 |
| Treatment/medication code: atenolol | 5.40E-04 | -0.0017 |
| FreeSurfer volume Left-Hippocampus | 5.70E-04 | -0.042 |
| Treatment/medication code: sotalol | 6.70E-04 | -0.00034 |
| Waist circumference | 7.00E-04 | 0.0074 |
| Treatment/medication code: otomize ear spray | 7.10E-04 | -0.00019 |
| Diastolic blood pressure, automated reading | 7.90E-04 | -0.0084 |
| Non-cancer illness code, self-reported: cystitis | 9.60E-04 | -0.00023 |
| Body mass index (BMI) | 1.00E-03 | 0.008 |
| T1 FAST ROIs L hippocampus | 1.30E-03 | -0.039 |
| FreeSurfer a2009s rh S circular insula ant area | 1.40E-03 | -0.045 |
| Weight | 1.40E-03 | 0.0069 |
| FreeSurfer volume Right-Hippocampus | 1.60E-03 | -0.039 |
| Non-cancer illness code, self-reported: urticaria | 1.60E-03 | -0.00019 |
| Triglycerides in small VLDL | 1.60E-03 | -0.036 |
| Treatment/medication code: minims artificial tears single-use eye drops | 1.70E-03 | -0.00011 |

<table>
<tr><th colspan="3">Open Target Genetics</th></tr>
<tr><th colspan="3">rs201632637 (proxy used: rs7633256, R2 = 0.94, Ref = CEU) (Nearest gene: KLF14)</th></tr>
<tr><th>Trait</th><th>P-value</th><th>Beta</th></tr>
<tr><td>Hip circumference</td><td>1.1114E-23</td><td>0.218279</td></tr>
<tr><td>Trunk fat percentage</td><td>1.17149E-15</td><td>0.138282</td></tr>
<tr><td>Body fat percentage</td><td>3.41403E-15</td><td>0.119041</td></tr>
<tr><td>Trunk fat mass</td><td>1.37821E-13</td><td>0.0911783</td></tr>
<tr><td>Arm fat percentage (right)</td><td>1.63832E-13</td><td>0.131641</td></tr>
<tr><td>Arm fat percentage (left)</td><td>1.72479E-13</td><td>0.132689</td></tr>
<tr><td>Blood pressure medication | medication for cholesterol, blood pressure, diabetes, or take e</td><td>2.56015E-13</td><td>-0.060737312</td></tr>
<tr><td>Cholesterol lowering medication | medication for cholesterol, blood pressure, diabetes, or</td><td>5.14072E-12</td><td>-0.065557865</td></tr>
<tr><td>Whole body fat mass</td><td>1.46556E-11</td><td>0.149246</td></tr>
<tr><td>Hypertension | non-cancer illness code, self-reported</td><td>2.67973E-11</td><td>-0.035159319</td></tr>
<tr><td>High blood pressure | vascular/heart problems diagnosed by doctor</td><td>5.76962E-11</td><td>-0.034223722</td></tr>
<tr><td>Leg fat percentage (right)</td><td>9.70148E-11</td><td>0.0851131</td></tr>
<tr><td>None of the above | vascular/heart problems diagnosed by doctor</td><td>1.21042E-10</td><td>0.032365939</td></tr>
<tr><td>Leg fat percentage (left)</td><td>3.82497E-10</td><td>0.0782785</td></tr>
<tr><td>Ankle spacing width</td><td>3.92828E-10</td><td>-0.0778254</td></tr>
<tr><td>Impedance of arm (right)</td><td>8.03923E-10</td><td>0.560406</td></tr>
<tr><td>Arm fat mass (left)</td><td>1.11134E-09</td><td>0.0100205</td></tr>
<tr><td>Arm fat mass (right)</td><td>1.87423E-09</td><td>0.00884281</td></tr>
<tr><td>Impedance of arm (left)</td><td>2.19311E-09</td><td>0.56</td></tr>
<tr><td>Leg fat mass (right)</td><td>4.83026E-08</td><td>0.0204372</td></tr>
<tr><td>None of the above | medication for cholesterol, blood pressure, diabetes, or take exogenou</td><td>9.98317E-08</td><td>0.036743182</td></tr>
<tr><td>Leg fat mass (left)</td><td>1.06501E-07</td><td>0.0194257</td></tr>
<tr><td>Simvastatin | treatment/medication code</td><td>3.86373E-07</td><td>-0.036973125</td></tr>
<tr><td>Impedance of whole body</td><td>5.0655E-07</td><td>0.800828</td></tr>
<tr><td>Mean platelet (thrombocyte) volume</td><td>5.54865E-07</td><td>0.0130192</td></tr>
<tr><td>Waist circumference</td><td>1.23602E-06</td><td>0.137289</td></tr>
<tr><td>Diabetes diagnosed by doctor</td><td>1.41716E-06</td><td>-0.053431076</td></tr>
<tr><td>High cholesterol | non-cancer illness code, self-reported</td><td>1.50076E-06</td><td>-0.034160174</td></tr>
<tr><td>Body mass index (bmi)</td><td>2.7714E-06</td><td>0.0528337</td></tr>
<tr><td>Body mass index (bmi)</td><td>3.15436E-06</td><td>0.0527417</td></tr>
<tr><td>Mean sphered cell volume</td><td>3.64165E-06</td><td>0.0590453</td></tr>
<tr><td>High light scatter reticulocyte count</td><td>9.18807E-06</td><td>-0.000110148</td></tr>
<tr><td>Metformin | treatment/medication code</td><td>1.98172E-05</td><td>-0.065028318</td></tr>
<tr><td>Type 2 diabetes</td><td>0.00002027</td><td>-0.0673</td></tr>
<tr><td>Lisinopril | treatment/medication code</td><td>2.15872E-05</td><td>-0.061456933</td></tr>
<tr><td>Weight</td><td>3.76943E-05</td><td>0.138848</td></tr>
<tr><td>Weight</td><td>3.91857E-05</td><td>0.137957</td></tr>
<tr><td>Essential hypertension</td><td>0.0000422</td><td>-0.0266</td></tr>
<tr><td>Diabetes | illnesses of mother</td><td>5.21121E-05</td><td>-0.034121237</td></tr>
<tr><td>Hypertension</td><td>5.83E-05</td><td>-0.026</td></tr>
<tr><td>Diabetes mellitus</td><td>0.0000878</td><td>-0.0427</td></tr>
<tr><td>Mean platelet volume</td><td>0.00009192</td><td>0.01433537</td></tr>
<tr><td>Comparative height size at age 10</td><td>9.85883E-05</td><td>0.00631378</td></tr>
<tr><td>Age when periods started (menarche)</td><td>0.000105668</td><td>-0.0093532</td></tr>
<tr><td>Reticulocyte count</td><td>0.0001362</td><td>-0.0138985</td></tr>
<tr><td>Type 2 diabetes</td><td>0.000139</td><td>-0.0429</td></tr>
<tr><td>Diabetes | non-cancer illness code, self-reported</td><td>0.000146696</td><td>-0.046336892</td></tr>
<tr><td>Reticulocyte fraction of red cells</td><td>0.0002526</td><td>-0.01332348</td></tr>
<tr><td>Atenolol | treatment/medication code</td><td>0.000409211</td><td>-0.043292271</td></tr>
<tr><td>Ulceration of intestine</td><td>0.00043</td><td>-0.195</td></tr>
</table>

**Supplementary Table 6:** Phenome-wide association study of rs8126001 at *RGS19-OPRL1*

| Oxford Brain Imaging Genetics (BIG) | | |
|---|---|---|
| **rs8126001 (Nearest gene: RGS19, OPRL1)** | | |
| **Phenotype** | **P-value** | **Beta** |
| Non-cancer illness code, self-reported: high cholesterol | 5.20E-12 | -0.0055 |
| Vascular/heart problems diagnosed by doctor: None of the above | 2.50E-11 | 0.0074 |
| Medication for cholesterol, blood pressure or diabetes: None of the above | 3.00E-10 | 0.011 |
| Medication for cholesterol, blood pressure or diabetes: Cholesterol lowering medication | 6.50E-10 | -0.0094 |
| Systolic blood pressure, automated reading | 3.70E-09 | -0.015 |
| Vascular/heart problems diagnosed by doctor: High blood pressure | 8.90E-09 | -0.0062 |
| Treatment/medication code: simvastatin | 3.00E-08 | -0.0043 |
| Medication for cholesterol, blood pressure, diabetes, or take exogenous hormones: Cholesterol lowering medication | 3.20E-08 | -0.0061 |
| Non-cancer illness code, self-reported: hypertension | 4.00E-08 | -0.0059 |
| Impedance of whole body | 1.90E-07 | -0.0097 |
| Number of self-reported non-cancer illnesses | 1.20E-06 | -0.0098 |
| Impedance of arm (left) | 3.30E-06 | -0.008 |
| Treatment/medication code: ramipril | 6.00E-06 | -0.0023 |
| Treatment/medication code: aspirin | 6.70E-06 | -0.0037 |
| Birth weight of first child | 7.40E-06 | 0.02 |
| Diagnoses - main ICD10: I25 Chronic ischaemic heart disease | 8.00E-06 | -0.0017 |
| dMRI TBSS ISOVF Superior corona radiata R | 8.10E-06 | 0.056 |
| Impedance of arm (right) | 9.60E-06 | -0.0076 |
| Medication for pain relief, constipation, heartburn: Aspirin | 1.10E-05 | -0.0037 |
| Illnesses of father: Heart disease | 1.10E-05 | -0.0054 |
| Impedance of leg (left) | 1.30E-05 | -0.0097 |
| Vascular/heart problems diagnosed by doctor: Angina | 2.80E-05 | -0.0018 |
| Impedance of leg (right) | 3.70E-05 | -0.0091 |
| Non-cancer illness code, self-reported: angina | 4.30E-05 | -0.0017 |
| dMRI ProbtrackX L1 slf l | 6.60E-05 | 0.055 |
| Medication for cholesterol, blood pressure, diabetes, or take exogenous hormones: Blood pressure medication | 7.80E-05 | -0.005 |
| Hip circumference | 8.10E-05 | 0.0095 |
| dMRI TBSS ISOVF Superior corona radiata L | 9.10E-05 | 0.051 |
| dMRI ProbtrackX ISOVF str l | 9.20E-05 | 0.049 |
| Medication for cholesterol, blood pressure or diabetes: Blood pressure medication | 9.30E-05 | -0.0061 |
| Medication for cholesterol, blood pressure, diabetes, or take exogenous hormones: None of the above | 1.10E-04 | 0.006 |
| Illnesses of mother: Diabetes | 1.10E-04 | -0.0028 |
| dMRI TBSS ISOVF Posterior thalamic radiation L | 1.40E-04 | 0.059 |
| Forced expiratory volume in 1-second (FEV1), predicted percentage | 1.40E-04 | 0.016 |
| Forced expiratory volume in 1-second (FEV1) | 1.50E-04 | 0.0079 |
| Had other major operations | 1.70E-04 | 0.0059 |
| Forced vital capacity (FVC) | 1.80E-04 | 0.0074 |
| Underlying (primary) cause of death: ICD10: C85.9 Non-Hodgkin's lymphoma, unspecified type | 1.80E-04 | -0.0058 |
| Treatment/medication code: atorvastatin | 2.00E-04 | -0.0016 |
| dMRI ProbtrackX ISOVF slf r | 2.30E-04 | 0.049 |
| dMRI ProbtrackX MD slf l | 3.20E-04 | 0.053 |
| dMRI ProbtrackX MD slf r | 3.20E-04 | 0.051 |
| dMRI ProbtrackX L1 slf r | 3.50E-04 | 0.047 |
| dMRI ProbtrackX L2 slf r | 3.70E-04 | 0.052 |
| Treatment/medication code: lisinopril | 3.90E-04 | -0.0014 |
| dMRI ProbtrackX ISOVF str r | 4.30E-04 | 0.042 |
| Diabetes diagnosed by doctor | 4.60E-04 | -0.0018 |
| Bring up phlegm/sputum/mucus on most days | 5.70E-04 | 0.0047 |
| dMRI TBSS MD Superior longitudinal fasciculus R | 5.90E-04 | 0.049 |
| FreeSurfer volume Left-Amygdala | 6.20E-04 | -0.042 |

| Open Target Genetics | | |
|---|---|---|
| **rs8126001 (Nearest gene: RGS19, OPRL1)** | | |
| **Trait** | **P-value** | **Beta** |
| High cholesterol \| non-cancer illness code, self-reported | 2.98385E-12 | -0.049164855 |
| None of the above \| vascular/heart problems diagnosed by doctor | 1.63862E-11 | 0.033599967 |
| Mean sphered cell volume | 1.29303E-10 | 0.0813431 |
| Mean corpuscular volume | 1.82902E-10 | 0.0664817 |
| None of the above \| medication for cholesterol, blood pressure or diabetes | 1.02225E-09 | 0.042516541 |
| Cholesterol lowering medication \| medication for cholesterol, blood pressure or diabetes | 1.4284E-09 | -0.047845461 |
| Cholesterol lowering medication \| medication for cholesterol, blood pressure, diabetes, or t | 2.7783E-09 | -0.056111864 |
| High blood pressure \| vascular/heart problems diagnosed by doctor | 4.8856E-09 | -0.030339626 |
| Mean corpuscular volume | 7.872E-09 | 0.02044855 |
| Hypertension \| non-cancer illness code, self-reported | 1.83693E-08 | -0.029460906 |
| Simvastatin \| treatment/medication code | 4.66822E-08 | -0.039491362 |
| Impedance of whole body | 5.06793E-08 | -0.861666 |
| Mean corpuscular haemoglobin | 5.70243E-07 | 0.0217278 |
| Ulcerative colitis | 7.042E-07 | 0.0824 |
| Mean corpuscular hemoglobin | 8.628E-07 | 0.01748823 |
| Ramipril \| treatment/medication code | 1.52901E-06 | -0.053328843 |
| Aspirin \| treatment/medication code | 1.72768E-06 | -0.032217619 |
| Angina \| vascular/heart problems diagnosed by doctor | 1.89807E-06 | -0.063614196 |
| Impedance of leg (right) | 2.45843E-06 | -0.361301 |
| Impedance of leg (left) | 2.59847E-06 | -0.359993 |
| Angina \| non-cancer illness code, self-reported | 3.36189E-06 | -0.062053629 |
| Impedance of arm (left) | 4.34686E-06 | -0.426591 |
| Aspirin \| medication for pain relief, constipation, heartburn | 4.67975E-06 | -0.030308392 |
| Heart disease \| illnesses of father | 4.86502E-06 | -0.024433554 |
| Number of self-reported non-cancer illnesses | 5.39996E-06 | -0.00859202 |
| Coronary artery disease | 0.000006469 | -0.0267 |
| Impedance of arm (right) | 7.80188E-06 | -0.404516 |
| Mean reticulocyte volume | 8.93096E-06 | 0.0830717 |
| Birth weight of first child | 1.41029E-05 | 0.0121927 |
| Coronary atherosclerosis | 0.0000156 | -0.0474 |
| Breast cancer | 0.00001752 | 0.0281 |
| Hot drink temperature | 3.85072E-05 | -0.00554822 |
| Blood pressure medication \| medication for cholesterol, blood pressure, diabetes, or take e | 4.06689E-05 | -0.033858793 |
| Atorvastatin \| treatment/medication code | 4.96798E-05 | -0.055829483 |
| Diabetes \| illnesses of mother | 5.04982E-05 | -0.033915878 |
| None of the above \| medication for cholesterol, blood pressure, diabetes, or take exogenou | 6.11986E-05 | 0.02747412 |
| Hip circumference | 7.29664E-05 | 0.0856447 |
| Hyperlipidemia | 7.51E-05 | -0.0331 |
| Coronary artery disease | 0.000076 | -0.027644 |
| Disorders of lipoid metabolism | 0.0000831 | -0.0329 |
| Inflammatory bowel disease | 0.0001192 | 0.0503 |
| Diabetes diagnosed by doctor | 0.000136592 | -0.041935756 |
| Forced expiratory volume in 1-second (fev1) | 0.000172968 | 0.00532793 |
| Blood pressure medication \| medication for cholesterol, blood pressure or diabetes | 0.000173131 | -0.02906805 |
| Place of birth in uk - north co-ordinate | 0.000204542 | 1063.93 |
| Eosinophill count | 0.000206532 | -0.00654894 |
| Forced vital capacity (fvc) | 0.000242762 | 0.00675107 |
| Had other major operations | 0.000278608 | 0.024880034 |
| Getting up in morning | 0.000292617 | 0.00643989 |
| Breast disorder nos | 0.000318 | 0.358 |

**Supplementary Table 7:** Phenome-wide association study of rs998584 at *VEGFA*

| Oxford Brain Imaging Genetics (BIG) | | |
|---|---|---|
| rs998584 (Nearest gene: *VEGFA* ) | | |
| **Phenotype** | **P-value** | **Beta** |
| Waist-to-hip ratio adjusted for BMI | 1.50E-18 | -0.037 |
| Hip circumference | 2.00E-18 | -0.021 |
| Waist circumference adjusted for BMI | 2.90E-14 | -0.031 |
| Arm fat percentage (left) | 7.30E-09 | -0.011 |
| Arm fat percentage (right) | 8.70E-09 | -0.011 |
| Non-cancer illness code, self-reported: high cholesterol | 3.50E-08 | 0.0044 |
| Impedance of leg (left) | 1.50E-07 | 0.012 |
| Trunk fat percentage | 1.50E-07 | -0.012 |
| Arm fat mass (right) | 2.70E-07 | -0.012 |
| Impedance of leg (right) | 3.20E-07 | 0.011 |
| Arm fat mass (left) | 3.20E-07 | -0.012 |
| Treatment/medication code: metformin | 6.90E-07 | 0.0019 |
| Trunk fat mass | 8.80E-07 | -0.012 |
| Vascular/heart problems diagnosed by doctor: None of the above | 1.30E-06 | -0.0054 |
| Leg predicted mass (left) | 2.70E-06 | -0.0075 |
| Leg fat-free mass (left) | 2.90E-06 | -0.0075 |
| Treatment/medication code: simvastatin | 3.10E-06 | 0.0036 |
| Vascular/heart problems diagnosed by doctor: High blood pressure | 3.70E-06 | 0.005 |
| Body mass index (BMI) | 4.30E-06 | -0.011 |
| Body fat percentage | 5.00E-06 | -0.0085 |
| Whole body fat mass | 5.10E-06 | -0.011 |
| Medication for cholesterol, blood pressure, diabetes, or take exogenous hormones: None of the above | 5.60E-06 | -0.007 |
| Concentration of large HDL particles | 7.60E-06 | -0.049 |
| Total lipids in large HDL | 8.20E-06 | -0.049 |
| Phospholipids in large HDL | 9.10E-06 | -0.049 |
| Non-cancer illness code, self-reported: hypertension | 9.20E-06 | 0.0047 |
| Weight | 1.20E-05 | -0.0093 |
| Leg fat-free mass (right) | 1.20E-05 | -0.007 |
| Leg predicted mass (right) | 1.30E-05 | -0.0069 |
| Total cholesterol in large HDL | 1.30E-05 | -0.048 |
| Cholesterol esters in large HDL | 1.50E-05 | -0.048 |
| Diabetes diagnosed by doctor | 1.80E-05 | 0.0022 |
| Medication for cholesterol, blood pressure, diabetes, or take exogenous hormones: Cholesterol lowering medication | 1.80E-05 | 0.0048 |
| Cholesterol esters in large VLDL | 1.80E-05 | 0.048 |
| Medication for cholesterol, blood pressure or diabetes: None of the above | 2.50E-05 | -0.0072 |
| Free cholesterol in large HDL | 2.50E-05 | -0.046 |
| Waist circumference | 3.90E-05 | 0.0089 |
| Hair/balding pattern: Pattern 4 | 4.40E-05 | 0.0057 |
| Pulse rate, automated reading | 5.30E-05 | 0.01 |
| Triglycerides in small VLDL | 5.60E-05 | 0.044 |
| Triglycerides in medium VLDL | 5.80E-05 | 0.044 |
| Illnesses of mother: Diabetes | 7.30E-05 | 0.0029 |
| Medication for cholesterol, blood pressure or diabetes: Cholesterol lowering medication | 8.20E-05 | 0.006 |
| Total lipids in large VLDL | 8.30E-05 | 0.043 |
| Triglycerides in large VLDL | 8.40E-05 | 0.043 |
| Phospholipids in medium VLDL | 9.90E-05 | 0.043 |
| Leg fat mass (right) | 1.10E-04 | -0.0075 |
| Free cholesterol in medium VLDL | 1.30E-04 | 0.042 |
| Medication for cholesterol, blood pressure or diabetes: Blood pressure medication | 1.40E-04 | 0.0059 |
| Total lipids in medium VLDL | 1.40E-04 | 0.042 |

<table>
<tr><th colspan="3">Open Target Genetics</th></tr>
<tr><th colspan="3">rs998584 (Nearest gene: VEGFA )</th></tr>
<tr><th>Trait</th><th>P-value</th><th>Beta</th></tr>
<tr><td>Hip circumference</td><td>7.03663E-19</td><td>-0.191518</td></tr>
<tr><td>Ankle spacing width</td><td>5.75557E-16</td><td>-0.0997339</td></tr>
<tr><td>Triglycerides</td><td>3.424E-15</td><td>0.0293</td></tr>
<tr><td>Reticulocyte count</td><td>2.524E-13</td><td>0.0263027</td></tr>
<tr><td>High light scatter reticulocyte count</td><td>6.235E-13</td><td>0.02581551</td></tr>
<tr><td>Red blood cell (erythrocyte) count</td><td>1.26146E-12</td><td>0.00593366</td></tr>
<tr><td>Mean sphered cell volume</td><td>1.38565E-12</td><td>-0.0896141</td></tr>
<tr><td>Haemoglobin concentration</td><td>6.69302E-12</td><td>0.0160456</td></tr>
<tr><td>Arm fat percentage (left)</td><td>2.25342E-11</td><td>-0.119418</td></tr>
<tr><td>Arm fat percentage (right)</td><td>7.44547E-11</td><td>-0.115231</td></tr>
<tr><td>High light scatter reticulocyte percentage of red cells</td><td>9.871E-11</td><td>0.02320853</td></tr>
<tr><td>Reticulocyte fraction of red cells</td><td>1.086E-10</td><td>0.02318643</td></tr>
<tr><td>Impedance of leg (left)</td><td>2.10582E-10</td><td>0.486331</td></tr>
<tr><td>Heel bone mineral density</td><td>2.4E-10</td><td>0.0129599</td></tr>
<tr><td>Impedance of leg (right)</td><td>2.45209E-10</td><td>0.485132</td></tr>
<tr><td>Platelet distribution width</td><td>3.46028E-10</td><td>0.0077039</td></tr>
<tr><td>High cholesterol | non-cancer illness code, self-reported</td><td>8.991E-10</td><td>0.043143918</td></tr>
<tr><td>Coronary artery disease</td><td>1.214E-09</td><td>0.0332</td></tr>
<tr><td>White blood cell count</td><td>6.616E-09</td><td>0.02080482</td></tr>
<tr><td>Trunk fat percentage</td><td>6.69347E-09</td><td>-0.0992862</td></tr>
<tr><td>Trunk fat mass</td><td>6.85967E-09</td><td>-0.0708034</td></tr>
<tr><td>High light scatter reticulocyte count</td><td>7.09405E-09</td><td>0.000142595</td></tr>
<tr><td>Myeloid white cell count</td><td>7.175E-09</td><td>0.02084357</td></tr>
<tr><td>Ankle spacing width (right)</td><td>9.97228E-09</td><td>-0.0951295</td></tr>
<tr><td>Hemoglobin concentration</td><td>1.06E-08</td><td>0.0203067</td></tr>
<tr><td>Sum neutrophil eosinophil counts</td><td>1.18E-08</td><td>0.02045818</td></tr>
<tr><td>Neutrophil count</td><td>1.248E-08</td><td>0.02040576</td></tr>
<tr><td>Sum basophil neutrophil counts</td><td>1.361E-08</td><td>0.02038575</td></tr>
<tr><td>Granulocyte count</td><td>1.371E-08</td><td>0.02039721</td></tr>
<tr><td>Coronary artery disease</td><td>0.000000025</td><td>0.0390679</td></tr>
<tr><td>Leg predicted mass (left)</td><td>2.61136E-08</td><td>-0.0149742</td></tr>
<tr><td>Leg fat-free mass (left)</td><td>3.27513E-08</td><td>-0.0158563</td></tr>
<tr><td>Haematocrit percentage</td><td>5.09843E-08</td><td>0.0375507</td></tr>
<tr><td>Leg fat-free mass (right)</td><td>6.98652E-08</td><td>-0.0154354</td></tr>
<tr><td>Leg predicted mass (right)</td><td>8.15378E-08</td><td>-0.0143983</td></tr>
<tr><td>Body mass index (bmi)</td><td>8.66478E-08</td><td>-0.0598367</td></tr>
<tr><td>Arm fat mass (left)</td><td>1.15251E-07</td><td>-0.00864417</td></tr>
<tr><td>Weight</td><td>1.16889E-07</td><td>-0.176273</td></tr>
<tr><td>Whole body fat mass</td><td>1.43751E-07</td><td>-0.1153</td></tr>
<tr><td>Arm fat mass (right)</td><td>1.46107E-07</td><td>-0.00767191</td></tr>
<tr><td>Body fat percentage</td><td>1.8643E-07</td><td>-0.0781267</td></tr>
<tr><td>Cholesterol lowering medication | medication for cholesterol, blood pressur</td><td>3.61553E-07</td><td>0.047993285</td></tr>
<tr><td>None of the above | vascular/heart problems diagnosed by doctor</td><td>5.48669E-07</td><td>-0.024974663</td></tr>
<tr><td>Type 2 diabetes</td><td>0.000000703</td><td>0.0553</td></tr>
<tr><td>None of the above | medication for cholesterol, blood pressure, diabetes, or</td><td>7.56822E-07</td><td>-0.033876507</td></tr>
<tr><td>Hematocrit</td><td>9.018E-07</td><td>0.0173552</td></tr>
<tr><td>Body mass index (bmi)</td><td>9.9616E-07</td><td>-0.0548938</td></tr>
<tr><td>Metformin | treatment/medication code</td><td>1.4943E-06</td><td>0.072737761</td></tr>
<tr><td>Simvastatin | treatment/medication code</td><td>1.60006E-06</td><td>0.034668979</td></tr>
<tr><td>Hypertension</td><td>0.00000182</td><td>0.0306</td></tr>
</table>

**Supplementary Figure 1:** Phenotypic (A) and genetic (B) correlations among the seven traits in the UK Biobank.

**A.**

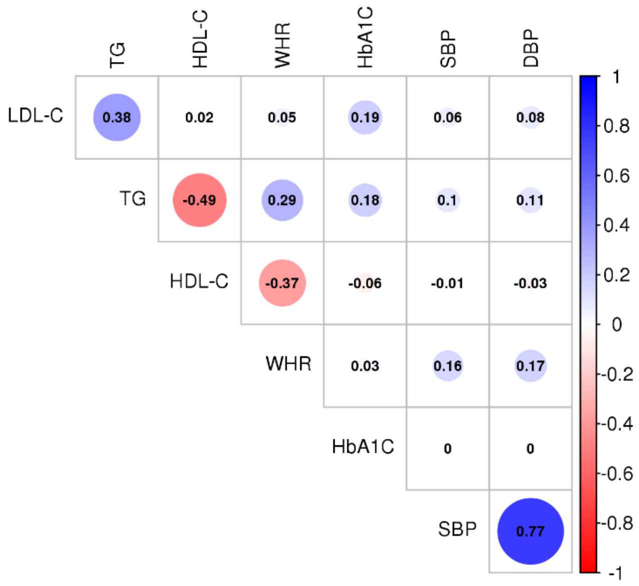


**B.**

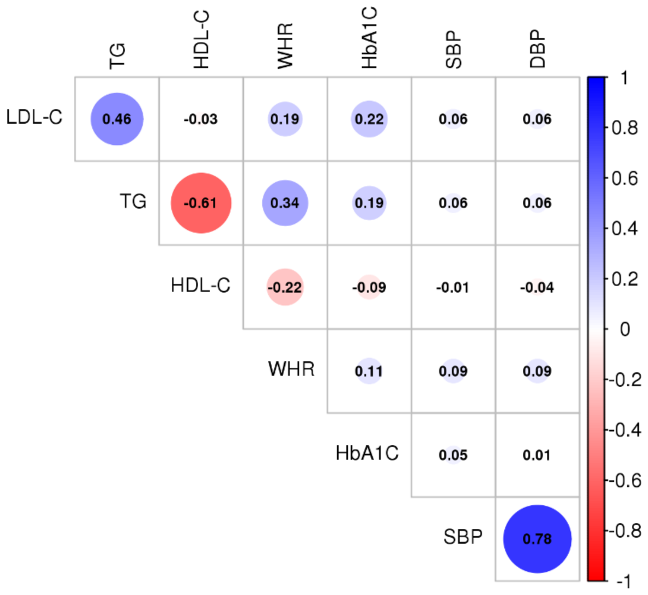
